# MDMA Assisted Psychotherapy Decreases PTSD Symptoms, Dissociation, Functional Disability, and Depression: A Systematic Review and Meta-Analysis

**DOI:** 10.1101/2023.08.17.23293955

**Authors:** W. M. Green, S.B. Raut, F.L.J. James, D.M. Benedek, R.J. Ursano, L.R. Johnson

## Abstract

Post-Traumatic Stress Disorder (PTSD) causes broad impairments affecting quality of life. However, despite current treatment many people with PTSD do not fully recover. MDMA assisted psychotherapy has emerged as a new therapy for PTSD and its comorbidities. We aimed to analyse the current evidence for MDMA assisted psychotherapy in PTSD and associated quality of life, and physiological effects, by conducting a systematic review and metanalysis of randomised controlled trials. ClinicalTrials.gov, MEDLINE, PsycINFO, PsycARTICLES, and Cochrane Library database were searched from inception to July 2022. We included both published and unpublished randomized control trials comparing MDMA assisted psychotherapy (MDMA-AP) with control. Meta-analysis of primary and secondary outcome measures was performed using Review-Manager software. Effect sizes were calculated using Standardised Mean Difference for CAPS scores and Mean Difference for secondary measures. MDMA-AP significantly improves dissociation, depression, and functional impairment, compared to controls, but not sleep quality. This data supports the use of MDMA-AP for PTSD with an improvement found in PTSD core symptoms and quality of life measures. While these findings are limited by small samples sizes in currently available clinical trials, this study provides empirical evidence to support development of MDMA-AP in PTSD.

## Introduction

Post-traumatic stress disorder (PTSD) is a devastating condition that develops in some individuals who fail to recover from exposure to extreme threatening situations, including actual or threatened death, serious injury, or sexual violence (Ursano et al., 2010). Symptoms include intrusive thoughts, avoidance, negative alteration in cognitions and mood, and marked alterations in arousal and reactivity (Merians, Spiller, Harpaz-Rotem, Krystal, & Pietrzak, 2023; Sanket B Raut, Marathe, et al., 2022a; Santiago et al., 2013; Ursano et al., 2010). Epidemiologic studies of international population data from 20 countries in a WHO World Mental Health Survey Initiative showed 12-month PTSD prevalence averaged 1.1%, ranging between 0.2-3.8% (Karam et al., 2014; Yehuda et al., 2015). While the aggregate lifetime prevalence of PTSD is estimated to be approximately 8% (Ahmadi et al., 2022; Kilpatrick et al., 2013; Lewis et al., 2019; Yehuda et al., 2015), the rate is substantially higher in certain populations. In United States veterans following combat in Iraq, prevalence rates vary from 9% to 31% (Dursa, Reinhard, Barth, & Schneiderman, 2014; Thomas et al., 2010). Despite its prevalence rate, unfortunately many patients do not recover from PTSD. Approximately one-fifth of patients develop an unremitting condition (Cusack et al., 2016; Fletcher, Creamer, & Forbes, 2010). Current treatment protocols include behavioural therapies (cognitive processing therapy and exposure therapy) usually over a 12-week period, and pharmacological therapies using SSRI and SNRI based treatments (fluoxetine, sertraline, paroxetine, and venlafaxine) these options do not provide complete recovery in all patients (Berger et al., 2009; John H. Krystal, Abdallah, et al., 2017; S. B. Raut, Marathe, et al., 2022b). Given the prevalence, impact, and lack of recovery in some patients, new and more successful treatment options for PTSD are urgently needed.

Psychedelics have been experimented with to assist psychotherapy since the 1970s and are currently ongoing a resurgence in interest for therapeutic use, including recent approval for use in PTSD by the Australian Therapeutic Goods Administration (TGA) (Bird, Modlin, & Rucker, 2021; Passie & Benzenhöfer, 2016; TGA, 2023). 3,4-Methylenedioxy methamphetamine (MDMA) was first synthesised in 1912 and is used as an illicit recreational drug commonly referred to as ‘ecstasy’. A lack of new treatments for PTSD has compelled renewed interest in psychedelic pharmacological research (John H Krystal, Davis, et al., 2017).

To date the Multidisciplinary Association for Psychedelic Studies (MAPS) is the only sponsor of global MDMA RCTs. MAPS have developed their own manualized MDMA assisted psychotherapeutic approach (MDMA-AP), involving 2 - 3 sessions of treatment (Emerson, Ponté, Jerome, & Doblin, 2014). A key element of the therapy is its non-directive approach (for full details see http://maps.org/treatment-manual). They hypothesised the pharmacokinetic action of MDMA, in combination with unique psychotherapy stimulates cognitive flexibility and reduces barriers to treating traumatic memories (Feduccia & Mithoefer, 2018; Gamma, Buck, Berthold, Hell, & Vollenweider, 2000). The mechanism of change is theorised to include memory reactivation, reconsolidation, and facilitated fear extinction (Feduccia & Mithoefer, 2018; S. B. Raut, Marathe, et al., 2022b). Previous meta-analyse find MDMA-AP reduces PTSD symptoms in treatment-resistant and severe PTSD populations and included analysis of safety and adverse events (Bahji, Forsyth, Groll, & Hawken, 2020; Hoskins et al., 2021; Illingworth et al., 2021; Michael C Mithoefer et al., 2019; Smith, Sicignano, Hernandez, & White, 2022; Tedesco et al., 2021). Data indicates the MDMA treatment effect is long-lasting (Bahji et al., 2020; Tedesco et al., 2021) and is effective in participants who are non-response to SSRI treatments (Tedesco et al., 2021). To date, meta-analyses have not incorporated measures of individual symptoms, comorbid conditions, and physiological responses. Here we hypothesize positive MDMA-AP treatment effects on PSTD associated dissociation, sleep quality, and functional disability, as well as comorbid depression.

## Methods

A systematic review and meta-analysis were conducted according to the Cochrane Intervention Review (MECIR) methodological expectation standards (J. Higgins, Lasserson, Chandler, Tovey, & Churchill, 2016), the American Psychological Association Journal Article Reporting Standards (JARS) Quant, Table 9 (APA, 2018), and the Preferred Reporting Items for Systematic Review and Meta-analysis (PRISMA) guidelines (Moher, Liberati, Tetzlaff, Altman, & Group*, 2009). The study protocol was registered with the International Prospective Register of Systematic Reviews (PROSPERO) on June 21, 2022. The registration identification number is CRD42022338904.

### Inclusion and Exclusion Criteria

We considered RCTs whose primary outcome variable was measured with the clinically administered PTSD scale (CAPS-IV, or CAPS-5) with either an active control or placebo comparator. Studies were included with adult participants diagnosed with PTSD. MDMA must have been clinically administered with psychotherapy to reduce PTSD symptoms. Comparison interventions were limited to psychotherapy alone. Studies were excluded if they were pre-clinical, animal, non-English, review articles, and infant or adolescent participants.

### Information Sources and Study Selection

MEDLINE, PsycINFO, PsycARTICLES, Cochrane Library database, and ClinicalTrials.gov were searched from inception until Friday, July 08, 2022. A specific approach was adopted using a highly sensitive search strategy (J. P. Higgins, Thomas, et al., 2019) for identifying RCTs indexed with associated medical subject headings (MeSH) terms (Baumann, 2016) only relating to or describing the population and intervention. The search strategy included the keywords 3, 4-Methylenedioxymethamphetamine, or MDMA, and, posttraumatic stress disorder, or PTSD. For example, (MDMA OR 3,4-methylenedioxymethamphetamine [MeSH Terms]) AND (PTSD OR Posttraumatic Stress Disorder [MeSH Terms]). Results were filtered where possible using the options, randomised controlled trials, humans, and English options.

Clinical trial registries were included in our search strategy to identify studies that have not been published in a journal to identify publication bias and selective reporting. Three researchers including one non-content expert, were assigned as reviewers to minimise review author bias. Two reviewers: (W.G., F.H.) screened the title and abstract, and selected studies were full text reviewed. Discrepancies were resolved by discussion. A third reviewer: (L.J.) resolved conflicts. Full texts were prepared for data extraction.

Six measures were identified for data collection and obtained after last treatment session. Effects of treatment were determined by using the difference in score following treatment. The following measures were used: (a) PTSD symptoms; CAPS, CAPS-IV, or CAPS-5, (b) comorbid depression; Beck’s Depression Inventory (BDI-II), (c) sleep disturbances; Pittsburgh Sleep Quality Index (PSQI), (d) dissociation severity; Disassociation Experience scale (DES-II), (e) functional disability; Global Assessment of Functioning scale (GAF), and Sheehan Disability Scale (SDS), (f) and physiological measures; heart rate, body temperature, systolic and diastolic blood pressure (S. B. Raut, Canales, et al., 2022). Two reviewers independently extracted the data (W.G., F.H.). The third reviewer resolved any inconsistencies.

### Assessing Risk to Internal Validity

Study quality was assessed following Cochranes’s risk of bias (RoB) items: (a) random sequence generation, (b) allocation concealment, (c) participant and personnel blinding, (d) outcome assessment blinding, (e) incomplete outcome data, (f) selective reporting, (g) other bias (J. P. Higgins, Savović, Page, Elbers, & Sterne, 2019). Two reviewers (W.G., F.J.) independently assessed the included study’s internal validity. To determine the risk, we checked if the included study authors reported the method used to address RoB in the journal or protocol publication. The quality assessment did not influence the decisions to synthesise any data.

### Summary Measures and Meta-Analytic Procedures

Where studies assessed the outcome of the dependent variable on the same measurement scale we combined the mean change score, standard deviation, and intention to treat sample size to calculate the mean difference (MD). Where the outcome of the dependent variable was assessed on different measurement scales, a standardised mean difference (SMD) model was used (J. P. Higgins, Thomas, et al., 2019; Murad, Wang, Chu, & Lin, 2019). The SMD is defined by Hedges (adjusted) g.

### Data Synthesis Methods

Covidence v2930, a web-based collaboration software program that streamlines the production of systematic reviews to administer the included studies was used with a customised template v2.0 to extract data. Only intention to treat participants were included. A meta-analysis for each primary and secondary outcome measure was performed using Review Manager v.5.4.1. We set a random effects model because it is assumed there is sample heterogeneity. We calculated 95% confidence intervals and two-tailedp-values. The between-study variance was estimated with Tau^2^. Heterogeneity was calculated with a chi-squared test producing an I^2^ statistic to assess the observed variance assuming all sampling errors could be removed. We considered an I^2^ value using the following thresholds: (a) 0% to 40%, might not be important; (b) 30% to 60%; may represent moderate heterogeneity, (c) 50% to 90%, may represent substantial heterogeneity; (d) 75% to 100%, considerable heterogeneity (J. P. Higgins, Thomas, et al., 2019). To measure the true variance, we calculated prediction intervals with the CMA prediction interval program using the MD (Borenstein, Higgins, Hedges, & Rothstein, 2017). In SMD models we used Cohan’s rule of thumb to interpret Hedges g: (a) 0.2 = small; (b) 0.5 = medium; (c) 0.8 = large.

## Results

### Study Selection and Characteristics

The study selection procedure is shown in **Figure 1**. 82 records were identified of which 27 reports sought for retrieval met the eligibility criteria. Six randomised psychotherapy-alone control comparisons were included for data extraction. Sample sizes ranged from 8 to 91 middle-aged adults. **Table 1 and 2** provide detailed characteristics and demographic summaries respectively. The complete data sets supporting this study are available from the study sponsor at maps.org/datause.

**Figure 1.**
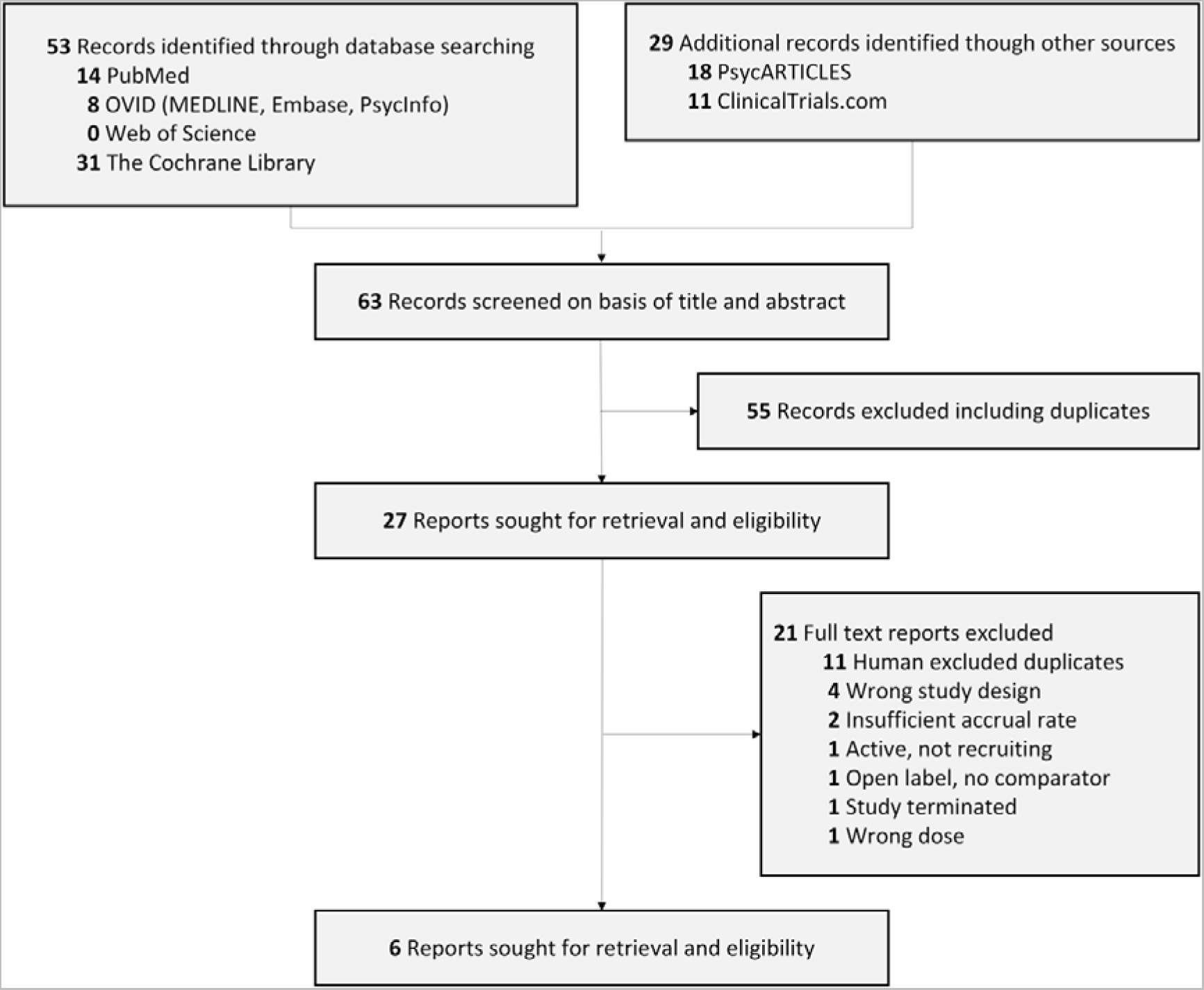
Study Selection Flow Chart.

**Table 1.**
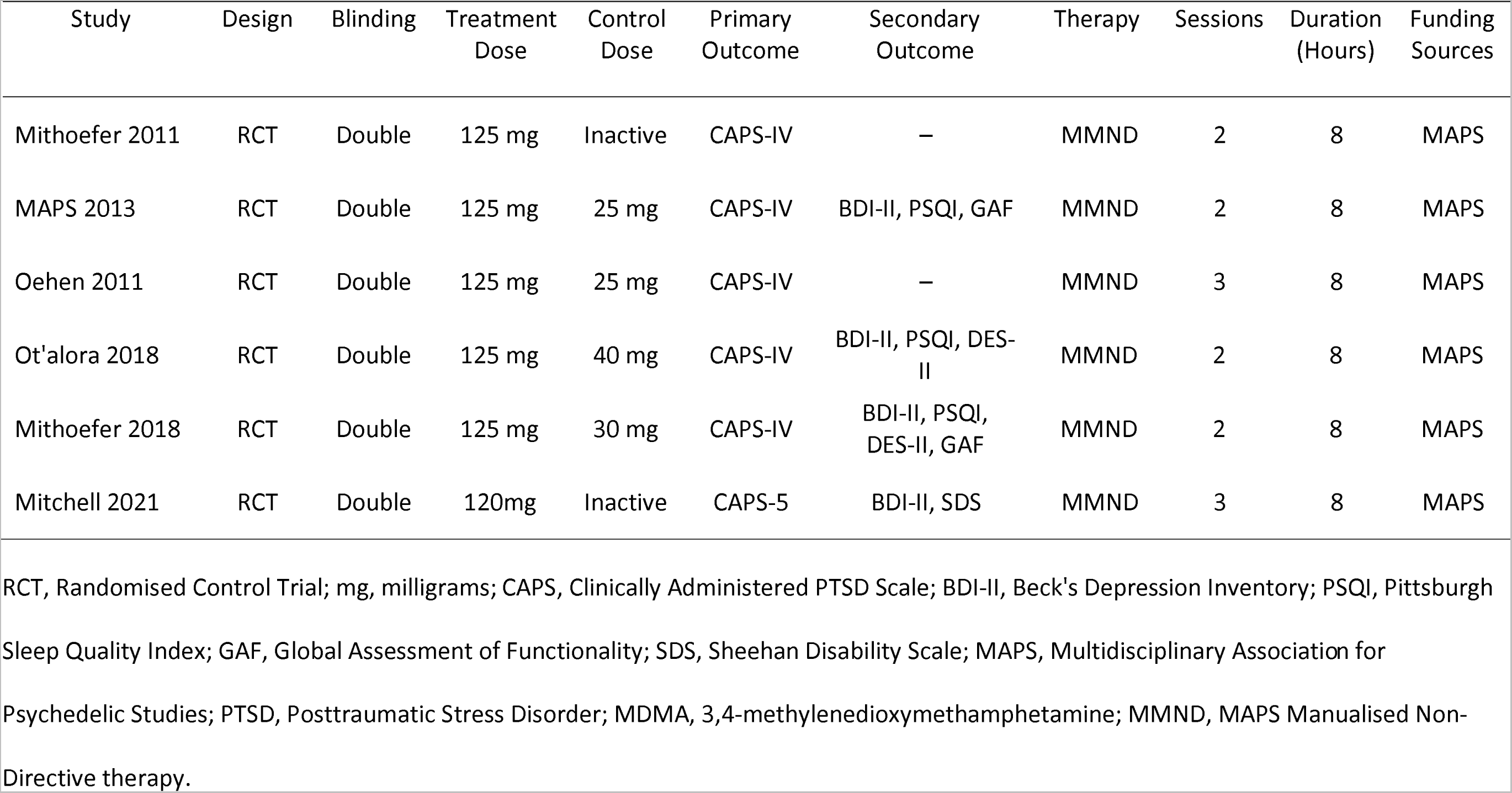
Study Characteristics for MDMA Treatment and Control Group.

**Table 2.** Study Demographics for MDMA Treatment and Control Group.

| Study | Randomised<br>*n* | Age |  | PTSD Duration |  | Gender (Treatment) |  | Gender (Control) |  |
| --- | --- | --- | --- | --- | --- | --- | --- | --- | --- |
|  |  | Treatment<br>*M* (SD) | Control<br>*M* (SD) | Treatment<br>*M* (SD) | Control<br>*M* (SD) | Male<br>*n* (%) | Female<br>*n* (%) | Male<br>*n* (%) | Female<br>*n* (%) |
| Mithoefer 2011 | 20 | 40.2 (7.6) | 40.8 (7.0) | 19.3 (16.8) | 22.8 (10.1) | 2 (17) | 10 (83) | 1 (13) | 7 (87) |
| MAPS 2013 | 8 | – | – | – | – | 2 (40) | 3 (60) | 3 (100) | 0 (0) |
| Oehen 2011 | 12 | 42.1 (12.8) | 40.0 (6.2) | 16.4 (10.9) | 22.3 (12.1) | 1 (12.5) | 7 (87.5) | 1 (25) | 3 (75) |
| Ot'alora 2018 | 19 | 44.6 (15.4) | 40.0 (11.7) | 33.9 (23.0) | 21.7 (13.6) | 5 (38.5) | 8 (61.5) | 1 (16.7) | 5 (83.3) |
| Mithoefer 2018 | 19 | 40.7 (11.1) | 39.2 (9.7) | 9.24 (7.1) | 5.7 (1.3) | 8 (67) | 4 (33) | 5 (71) | 2 (29) |
| Mitchell 2021 | 91 | 43.5 (12.9) | 38.2 (10.4) | 14.8 (11.6) | 13.2 (11.4) | 19 (41.3) | 27 (58.7) | 12 (27.3) | 32 (72.7) |

Age and duration are measured in years. Cells with a dash represent data not reported. PTSD, Posttraumatic Stress Disorder; MDMA, 3,4-methylenedioxymethamphetamine.

Preferred reporting items for systematic review and meta-analysis 2020 (PRISMA) for new systematic reviews, which included a register and database search. Adapted from Page et al. (2021).

### Effectiveness of MDMA-AP

A large effect of MDMA-AP occurring in 2-3 treatments sessions was observed on primary outcomes (PTSD symptoms: *g* = 0.93; 95% CI, 0.60 to 1.25; *p*<.001; PI, 0.52 to 1.38; n = 169). There was no significant amount of heterogeneity (Q(5) = 1.87; *p* = 0.87; tau^2^ = 0.00; *I*^2^ = 0.00%). For most secondary measures the outcome was significant and with similarly large effect sizes. For, independently measured, dissociation severity the mean difference (*MD) was* −9.7; 95% CI, −13.39 to −6.12; *p*<.001; n = 38. For, independently measured, daily functioning the standardised mean difference calculate effect size was *g* = 0.82; 95% CI, 0.10 to 1.73, *p =* 0.03; n = 118. For, independently measured, comorbid depression the mean difference (*MD) was,* −11.13; 95% CI, −19.35 to −2.92; *p* = 0.008; n = 137.

The observed result was smaller and not significant in sleep disturbance severity (*MD,* −3.67; 95% CI, −7.70 to 0.36; *p =* 0.07, n = 46). Of physiological measures heart rate was observed to increase (*MD,* 12.88; 95% CI, 0.97 to 24.79; *p* = 0.03; n = 32). Heterogeneity varied (dissociation severity: *I*^2^ = 7.00%; comorbid depression: *I*^2^ = 72.00%; functional disability: *I*^2^ = 52.00%; sleep disturbance: *I*^2^ = 65.00%; heart rate: *I*^2^ = 0.00%). **See Figure 2**.

**Figure 2.**
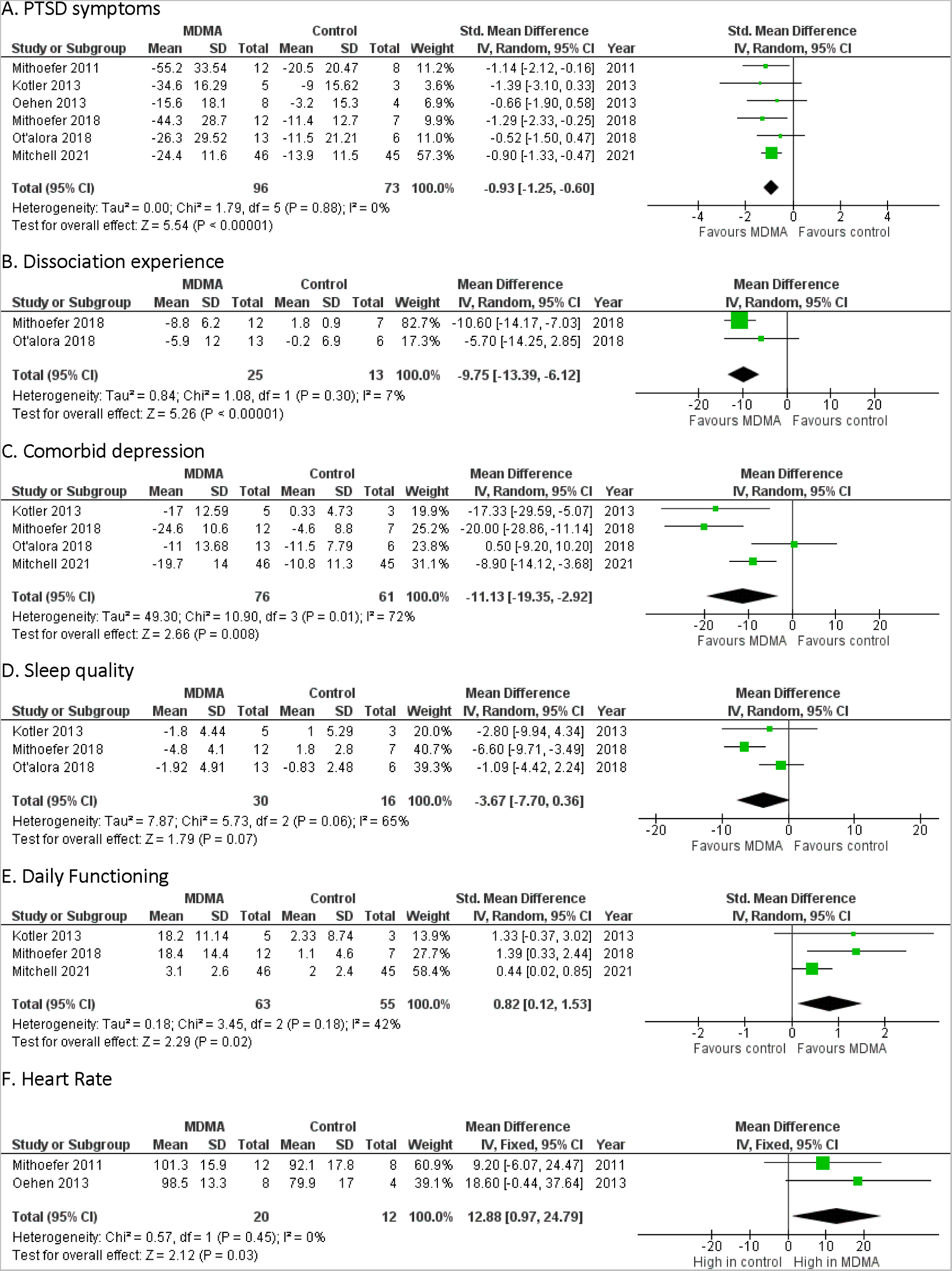
Effect of MDMA-Assisted Psychotherapy Compared to Psychotherapy Alone.

### Confidence in the Evidence

Internal validity of the included studies was moderate to high. Results were stratified by study quality, **see figure 3**. The same effect on the primary outcomes emerged with a small broadening of confidence intervals was observed after two high RoB studies were excluded from the analysis (*g* = 0.93; 95% CI, 1.27 to 0.58; *p*<.001). There is no evidence of publication bias measured with Eggars Regression (*p* = 0.07). One study not journal published was detected. However, MDMA use protocol requires clinical trials to be registered therefore there is no reason to believe other null or weaker studies exist.

**Figure 3.**
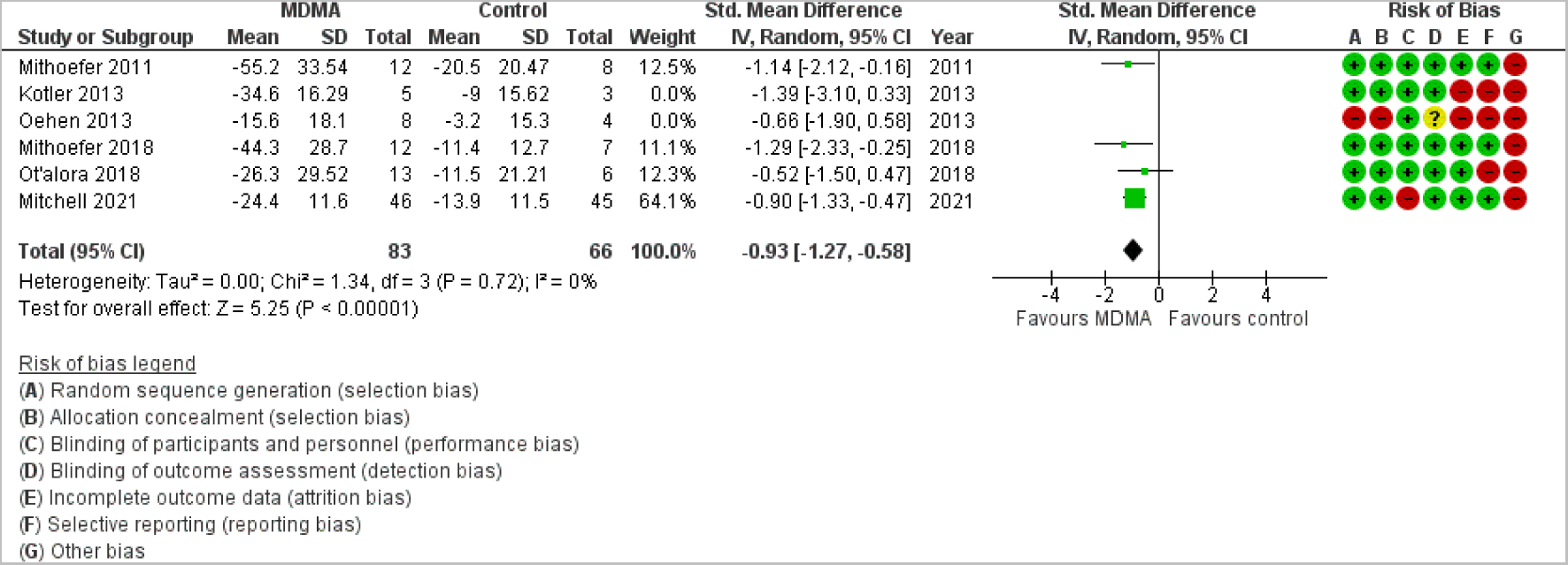
Stratified Results by Risk of Bias.

## Discussion

This meta-analysis is the most current and complete assessment of MDMA-AP in people diagnosed with severe or treatment resistant PTSD, synthesizing six RCTs conducted between 2011 and 2021. Importantly, we also include the first comprehensive analysis of secondary outcome measures and find MDMA-AP significantly improves dissociation, depression, and functional impairment, finding large effect sizes compared to controls, but not for sleep quality. This study is also the first meta-analysis to examine the effect of MDMA-AP in RCTs on physiological measures of heart rate, diastolic and systolic blood pressure, and body temperature, finding participants experience a significant increase in heart rate during treatment sessions. Together, these new results provide evidence for broad therapeutic advantages of MDMA-AP across the PTSD symptom profile.

The present meta-analysis finds MDMA-AP has effects on PTSD symptoms within a few treatment sessions. Traditional therapies including Cognitive Processing Therapy and SSRI treatments take weeks to months for symptom relief to occur (Gutner, Gallagher, Baker, Sloan, & Resick, 2016). In contrast, in the studies identified in this systematic review and meta-analysis we find MDMA-AP symptoms improvements can occur within 2-3 treatment sessions. Participants were previously unsuccessful using current treatment approaches however they reported a significant reduction in PTSD symptoms and comorbid conditions after MDMA-AP (Kotler, 2013; Mitchell et al., 2021; M. C. Mithoefer et al., 2018). Therapies with effects in minimal treatment sessions are likely to get more compliance with increased satisfaction. In addition to having rapid effects on PTSD symptoms measured with CAPS, we find MDMA-AP also acts on key secondary measures of PTSD in randomised clinical trials.

Dissociation is a hallmark of severe PTSD and is characterised by depersonalisation and derealisation symptoms. Our results indicate MDMA-AP treatment significantly reduces DES II scores compared to control. This suggests the treatment reduces dissociation severity and is likely because MDMA enhances the psychotherapeutic process that reduces the intense emotions associated with PTSD (Pai, Suris, & North, 2017). The result is clinically useful because almost half of PTSD diagnoses include dissociation incidents (White et al., 2022).

The current meta-analysis shows MDMA-AP reduces comorbid depression. Treatment guidelines for PTSD advise treatment efficacy is limited by severe depression (Martin et al., 2021). However, this result supports the rationale that psychedelics reduce both PTSD symptoms and comorbid depression (Bird et al., 2021; Michael C Mithoefer et al., 2019). The present meta-analysis includes two more RCTs and contradicts a previous meta-analysis by Illingworth et al. (2021) which showed MDMA-AP does not reduce depression. This result identifies a reduction in severe comorbid depression in PTSD.

Decreasing PTSD symptoms and comorbid conditions has been shown to improve cognitive capacity, daily functioning, and quality of life (Jellestad, Vital, Malamud, Taeymans, & Mueller-Pfeiffer, 2021). As individuals’ ability to mobilise the help they require increases, their health conditions decrease (Sareen, 2014). The present meta-analysis demonstrates participants treated with MDMA-AP experience improvements in daily functioning across psychological, social, and occupational domains compared to controls (Kotler, 2013; Mitchell et al., 2021; M. C. Mithoefer et al., 2018). Furthermore, following treatment, participants report their PTSD symptom improvement remains stable from one to 3.8 years (Ponte et al., 2021), implying a similar long-term improvement in daily functioning.

A cardinal feature of PTSD is sleep disturbance. Correspondingly, decreasing PTSD symptoms increases sleep quality to improve quality of life (de Moraes Costa et al., 2022; Maher et al., 2021). Our results indicate MDMA-AP reduces PQSI scores used to measure sleep quality however the reduction in score was not statistically significant (p = 0.07). Sleep quality is observed to improve at follow-up (Ponte et al., 2021). Perhaps the result was not significant because sleep disturbance is assessed at the primary endpoint when the acute effect of MDMA on participants is extant (Ot’alora et al., 2018; Parrott, 2013). PTSD symptom, comorbid condition and functional disability improvement may result in a delayed reduction in sleep disturbance (Slavish, Briggs, Fentem, Messman, & Contractor, 2022).

The meta-analysis showed MDMA-AP increases heart rate during treatment sessions but not diastolic blood pressure or body temperature. Results indicate the treatment’s acute effect increases heart rate by an average of 13 bpm compared to controls. This finding supports a relationship between cardiac-specific effects and the risk of cardiac dysfunction (Fonseca, Ribeiro, Tapadas, & Cotrim, 2021). MDMA is known to increase sympathetic nervous system activation and therefore increase cardiac vulnerability in PTSD (Edmondson & von Känel, 2017). The result does not challenge the intervention’s safety in people with treatment-resistant PTSD (Bahji et al., 2020). Instead, it suggests caution as future participants with underlying cardiac vulnerability may be at risk (Smith et al., 2022).

In addition to our findings on PTSD symptoms and secondary measures, this meta-analysis has several important strengths. In this study we have advanced our understanding of MDMA-AP by including the results of an unpublished RCT and our meta-analysis has allowed for variation in CAPS scales. While both the construct (PTSD) and the measure (CAPS) were consistent across studies, we identified that different studies used different versions of CAPS (DSM-IV versus DSM-5) with a different score range. While previous meta-analyses (Smith et al., 2022) have used the mean difference to compare RCT our use of the standardized mean difference approach accounts for version differences of CAPS.

The results of this study indicate benefits of MDMA-AP for the treatment of PTSD, however there are limitations of the findings of this meta-analysis. In its current form, MAPS non-directive psychotherapy has not been tested as a stand-alone therapy against established psychotherapies (for example, CPT, Trauma focused CBT, EMDR). Moreover, this therapy has not been independently standardised and replicated outside of MAPS and requires more research on PTSD patients worldwide. An additional limitation is the single study sponsor for all studies, highlighting the need for independent replication. An additional limitation of the findings is the small sample sizes that do not provide sufficient power to measure the secondary and primary outcomes in all studies except the single Phase 3 RCT.

## Conclusion

Evidence indicates that combining MDMA (120mg – 125mg) with psychotherapy to be very effective in reducing PTSD CAPS scores as well as significantly improving secondary PTSD measures and co-morbidities including dissociation, depression, quality of life and may also improve sleep quality. Together, this evidence provides a wide and comprehensive clinical picture for PTSD symptom improvement using MDMA-AP. Independent research groups need to assess the effect in more RCTs including comparisons with existing evidence-based therapies, existing FDA approved pharmacological treatments and in larger cohorts to increase the certainty of the findings. Most importantly, the type of psychotherapy practised in these trials is a substantial limiting factor in the clinical utility of this breakthrough treatment, a suitable evidence-based psychotherapy to combine with MDMA must be identified and evaluated in future trials.

## Data Availability

All data produced in the present work are contained in the manuscript

## Funding acknowledgement

We acknowledge University of Tasmania, College of Health and Medicine for funding and support.

## Conflict of Interest

The authors declare no conflict of Interest.

## Notes

### Competing Interest Statement

The authors have declared no competing interest.

### Author Declarations

The study used only openly available human data that were originally located at: 1. https://pubmed.ncbi.nlm.nih.gov/33972795/ 2.https://journals.sagepub.com/doi/full/10.1177/0269881112464827?rfr_dat=cr_pub++0pubmed&url_ver=Z39.88-2003&rfr_id=ori%3Arid%3Acrossref.org 3.https://www.sciencedirect.com/science/article/abs/pii/S2215036618301354?via%3Dihub 4.https://www.ncbi.nlm.nih.gov/pmc/articles/PMC3122379/ 5.https://www.ncbi.nlm.nih.gov/pmc/articles/PMC6247454/

## References

Ahmadi, R., Rahimi, S., Olfati, M., Javaheripour, N., Emamian, F., Ghadami, M. R., … Sepehry, A. A. (2022). Insomnia and post-traumatic stress disorder: A meta-analysis on interrelated association (n= 57,618) and prevalence (n= 573,665). Neuroscience & Biobehavioral Reviews, 104850.

APA. (2018). American Psychological Association, JARS Quant Table 9 Meta-Analysis Reporting Table.

Bahji, A., Forsyth, A., Groll, D., & Hawken, E. R. (2020). Efficacy of 3, 4-methylenedioxymethamphetamine (MDMA)-assisted psychotherapy for posttraumatic stress disorder: a systematic review and meta-analysis. Progress in Neuro-Psychopharmacology and Biological Psychiatry, 96, 109735.

Baumann, N. (2016). How to use the medical subject headings (MeSH). International journal of clinical practice, 70(2), 171–174.

Berger, W., Mendlowicz, M. V., Marques-Portella, C., Kinrys, G., Fontenelle, L. F., Marmar, C. R., & Figueira, I. (2009). Pharmacologic alternatives to antidepressants in posttraumatic stress disorder: a systematic review. Prog Neuropsychopharmacol Biol Psychiatry, 33(2), 169–180. doi:10.1016/j.pnpbp.2008.12.004

Bird, C. I., Modlin, N. L., & Rucker, J. J. (2021). Psilocybin and MDMA for the treatment of trauma-related psychopathology. International Review of Psychiatry, 33(3), 229–249.

Borenstein, M., Higgins, J. P., Hedges, L. V., & Rothstein, H. R. (2017). Basics of meta-analysis: I(2) is not an absolute measure of heterogeneity. Res Synth Methods, 8(1), 5–18. doi:10.1002/jrsm.1230

Cusack, K., Jonas, D. E., Forneris, C. A., Wines, C., Sonis, J., Middleton, J. C., … Gaynes, B. N. (2016). Psychological treatments for adults with posttraumatic stress disorder: A systematic review and meta-analysis. Clinical Psychology Review, 43, 128–141. https://doi.org/10.1016/j.cpr.2015.10.003

de Moraes Costa, G., Ziegelmann, P. K., Zanatta, F. B., Martins, C. C., de Moraes Costa, P., & Mello, C. F. (2022). Efficacy, acceptability, and tolerability of antidepressants for sleep quality disturbances in post-traumatic stress disorder: A systematic review and network meta-analysis. Progress in Neuro-Psychopharmacology and Biological Psychiatry, 110557.

Dursa, E. K., Reinhard, M. J., Barth, S. K., & Schneiderman, A. I. (2014). Prevalence of a positive screen for PTSD among OEF/OIF and OEF/OIF-era veterans in a large population-based cohort. J Trauma Stress, 27(5), 542–549. doi:10.1002/jts.21956

Edmondson, D., & von Känel, R. (2017). Post-traumatic stress disorder and cardiovascular disease. Lancet Psychiatry, 4(4), 320–329. doi:10.1016/s2215-0366(16)30377-7

Emerson, A., Ponté, L., Jerome, L., & Doblin, R. (2014). History and future of the Multidisciplinary Association for Psychedelic Studies (MAPS). Journal of psychoactive drugs, 46(1), 27–36.

Feduccia, A. A., & Mithoefer, M. C. (2018). MDMA-assisted psychotherapy for PTSD: Are memory reconsolidation and fear extinction underlying mechanisms?Prog Neuropsychopharmacol Biol Psychiatry, 84(Pt A), 221–228. doi:10.1016/j.pnpbp.2018.03.003

Fletcher, S., Creamer, M., & Forbes, D. (2010). Preventing post traumatic stress disorder: are drugs the answer? Aust N Z J Psychiatry, 44(12), 1064–1071. doi:10.3109/00048674.2010.509858

Fonseca, D. A., Ribeiro, D. M., Tapadas, M., & Cotrim, M. D. (2021). Ecstasy (3,4-methylenedioxymethamphetamine): Cardiovascular effects and mechanisms.Eur J Pharmacol, 903, 174156. doi:10.1016/j.ejphar.2021.174156

Gamma, A., Buck, A., Berthold, T., Hell, D., & Vollenweider, F. X. (2000). 3, 4-Methylenedioxymethamphetamine (MDMA) modulates cortical and limbic brain activity as measured by [H215O]-PET in healthy humans. Neuropsychopharmacology, 23(4), 388–395.

Gutner, C. A., Gallagher, M. W., Baker, A. S., Sloan, D. M., & Resick, P. A. (2016). Time course of treatment dropout in cognitive–behavioral therapies for posttraumatic stress disorder. *Psychological Trauma: Theory, Research*, Practice, and Policy, 8, 115–121. doi:10.1037/tra0000062

Higgins, J., Lasserson, T., Chandler, J., Tovey, D., & Churchill, R. (2016). Methodological expectations of Cochrane intervention reviews. London: Cochrane, 5.

Higgins, J. P., Savović, J., Page, M. J., Elbers, R. G., & Sterne, J. A. (2019). Assessing risk of bias in a randomized trial. Cochrane handbook for systematic reviews of interventions, 205–228.

Higgins, J. P., Thomas, J., Chandler, J., Cumpston, M., Li, T., Page, M. J., & Welch, V. A. (2019). Cochrane handbook for systematic reviews of interventions: John Wiley & Sons.

Hoskins, M. D., Sinnerton, R., Nakamura, A., Underwood, J. F., Slater, A., Lewis, C., … Clarke, L. (2021). Pharmacological-assisted Psychotherapy for Post Traumatic Stress Disorder: a systematic review and meta-analysis. European Journal of Psychotraumatology, 12(1), 1853379.

Illingworth, B. J., Lewis, D. J., Lambarth, A. T., Stocking, K., Duffy, J. M., Jelen, L. A., & Rucker, J. J. (2021). A comparison of MDMA-assisted psychotherapy to non-assisted psychotherapy in treatment-resistant PTSD: A systematic review and meta-analysis. Journal of Psychopharmacology, 35(5), 501–511.

Jellestad, L., Vital, N. A., Malamud, J., Taeymans, J., & Mueller-Pfeiffer, C. (2021). Functional impairment in posttraumatic stress disorder: a systematic review and meta-analysis. Journal of psychiatric research, 136, 14–22.

Karam, E. G., Friedman, M. J., Hill, E. D., Kessler, R. C., McLaughlin, K. A., Petukhova, M., … Bromet, E. J. (2014). Cumulative traumas and risk thresholds: 12-month PTSD in the World Mental Health (WMH) surveys. Depression and anxiety, 31(2), 130–142.

Kilpatrick, D. G., Resnick, H. S., Milanak, M. E., Miller, M. W., Keyes, K. M., & Friedman, M. J. (2013). National estimates of exposure to traumatic events and PTSD prevalence using DSM-IV and DSM-5 criteria. J Trauma Stress, 26(5), 537–547. doi:10.1002/jts.21848

Kotler, M. (2013). Randomized, Double-blind, Active Placebo-Controlled Pilot Study of MDMA-assisted Psychotherapy in People With Chronic PTSD. In. ClinicalTrials.gov identifier: NCT01689740.

Krystal, J. H., Abdallah, C. G., Averill, L. A., Kelmendi, B., Harpaz-Rotem, I., Sanacora, G., … Duman, R. S. (2017). Synaptic Loss and the Pathophysiology of PTSD: Implications for Ketamine as a Prototype Novel Therapeutic. Curr Psychiatry Rep, 19(10), 74. doi:10.1007/s11920-017-0829-z

Krystal, J. H., Davis, L. L., Neylan, T. C., Raskind, M. A., Schnurr, P. P., Stein, M. B., … Huang, G. D. (2017). It is time to address the crisis in the pharmacotherapy of posttraumatic stress disorder: a consensus statement of the PTSD Psychopharmacology Working Group. Biological Psychiatry, 82(7), e51–e59.

Lewis, S. J., Arseneault, L., Caspi, A., Fisher, H. L., Matthews, T., Moffitt, T. E., … Danese, A. (2019). The epidemiology of trauma and post-traumatic stress disorder in a representative cohort of young people in England and Wales. The Lancet Psychiatry, 6(3), 247–256. https://doi.org/10.1016/S2215-0366(19)30031-8

Maher, A. R., Apaydin, E. A., Hilton, L., Chen, C., Troxel, W., Hall, O., … Hempel, S. (2021). Sleep management in posttraumatic stress disorder: a systematic review and meta-analysis. Sleep Medicine, 87, 203–219.

Martin, A., Naunton, M., Kosari, S., Peterson, G., Thomas, J., & Christenson, J. K. (2021). Treatment Guidelines for PTSD: A Systematic Review. J Clin Med, 10(18). doi:10.3390/jcm10184175

Merians, A. N., Spiller, T., Harpaz-Rotem, I., Krystal, J. H., & Pietrzak, R. H. (2023). Post-traumatic Stress Disorder. Medical Clinics, 107(1), 85–99.

Mitchell, J. M., Bogenschutz, M., Lilienstein, A., Harrison, C., Kleiman, S., Parker-Guilbert, K., … Doblin, R. (2021). MDMA-assisted therapy for severe PTSD: a randomized, double-blind, placebo-controlled phase 3 study. Nat Med, 27(6), 1025–1033. doi:10.1038/s41591-021-01336-3

Mithoefer, M. C., Feduccia, A. A., Jerome, L., Mithoefer, A., Wagner, M., Walsh, Z., … Doblin, R. (2019). MDMA-assisted psychotherapy for treatment of PTSD: study design and rationale for phase 3 trials based on pooled analysis of six phase 2 randomized controlled trials. Psychopharmacology, 236(9), 2735–2745.

Mithoefer, M. C., Mithoefer, A. T., Feduccia, A. A., Jerome, L., Wagner, M., Wymer, J., … Doblin, R. (2018). 3,4-methylenedioxymethamphetamine (MDMA)-assisted psychotherapy for post-traumatic stress disorder in military veterans, firefighters, and police officers: a randomised, double-blind, dose-response, phase 2 clinical trial. Lancet Psychiatry, 5(6), 486–497. doi:10.1016/s2215-0366(18)30135-4

Moher, D., Liberati, A., Tetzlaff, J., Altman, D. G., & Group*, P. (2009). Preferred reporting items for systematic reviews and meta-analyses: the PRISMA statement.Annals of internal medicine, 151(4), 264–269.

Murad, M. H., Wang, Z., Chu, H., & Lin, L. (2019). When continuous outcomes are measured using different scales: guide for meta-analysis and interpretation. BMJ, 364, k4817. doi:10.1136/bmj.k4817

Ot’alora, G. M., Grigsby, J., Poulter, B., Van Derveer, J. W., 3rd, Giron, S. G., Jerome, L., … Doblin, R. (2018). 3,4-Methylenedioxymethamphetamine-assisted psychotherapy for treatment of chronic posttraumatic stress disorder: A randomized phase 2 controlled trial. J Psychopharmacol, 32(12), 1295–1307. doi:10.1177/0269881118806297

Page, M. J., McKenzie, J. E., Bossuyt, P. M., Boutron, I., Hoffmann, T. C., Mulrow, C. D., … Brennan, S. E. (2021). The PRISMA 2020 statement: an updated guideline for reporting systematic reviews. Systematic reviews, 10(1), 1–11.

Pai, A., Suris, A. M., & North, C. S. (2017). Posttraumatic Stress Disorder in the DSM-5: Controversy, Change, and Conceptual Considerations. Behav Sci (Basel), 7(1). doi:10.3390/bs7010007

Parrott, A. C. (2013). Human psychobiology of MDMA or ‘Ecstasy’: an overview of 25 years of empirical research. Human Psychopharmacology: Clinical and Experimental, 28(4), 289–307.

Passie, T., & Benzenhöfer, U. (2016). The History of MDMA as an Underground Drug in the United States, 1960-1979. J Psychoactive Drugs, 48(2), 67–75. doi:10.1080/02791072.2015.1128580

Ponte, L., Jerome, L., Hamilton, S., Mithoefer, M. C., Yazar-Klosinski, B. B., Vermetten, E., & Feduccia, A. A. (2021). Sleep Quality Improvements After MDMA-Assisted Psychotherapy for the Treatment of Posttraumatic Stress Disorder. Journal of traumatic stress, 34(4), 851–863.

Raut, S. B., Canales, J. J., Ravindran, M., Eri, R., Benedek, D. M., Ursano, R. J., & Johnson, L. R. (2022). Effects of propranolol on the modification of trauma memory reconsolidation in PTSD patients: A systematic review and meta-analysis. J Psychiatr Res, 150, 246–256. https://doi.org/10.1016/j.jpsychires.2022.03.045

Raut, S. B., Marathe, P. A., van Eijk, L., Eri, R., Ravindran, M., Benedek, D. M., … Johnson, L. R. (2022a). Diverse therapeutic developments for post-traumatic stress disorder (PTSD) indicate common mechanisms of memory modulation. Pharmacology & Therapeutics, 108195.

Raut, S. B., Marathe, P. A., van Eijk, L., Eri, R., Ravindran, M., Benedek, D. M., … Johnson, L. R. (2022b). Diverse therapeutic developments for post-traumatic stress disorder (PTSD) indicate common mechanisms of memory modulation. Pharmacol Ther, 239, 108195. https://doi.org/10.1016/j.pharmthera.2022.108195

Santiago, P. N., Ursano, R. J., Gray, C. L., Pynoos, R. S., Spiegel, D., Lewis-Fernandez, R., … Fullerton, C. S. (2013). A systematic review of PTSD prevalence and trajectories in DSM-5 defined trauma exposed populations: intentional and non-intentional traumatic events. PLoS One, 8(4), e59236. doi:10.1371/journal.pone.0059236

Sareen, J. (2014). Posttraumatic stress disorder in adults: impact, comorbidity, risk factors, and treatment. Can J Psychiatry, 59(9), 460–467. doi:10.1177/070674371405900902

Slavish, D. C., Briggs, M., Fentem, A., Messman, B. A., & Contractor, A. A. (2022). Bidirectional associations between daily PTSD symptoms and sleep disturbances: A systematic review. Sleep Med Rev, 63, 101623. doi:10.1016/j.smrv.2022.101623

Smith, K. W., Sicignano, D. J., Hernandez, A. V., & White, C. M. (2022). MDMA-assisted psychotherapy for treatment of posttraumatic stress disorder: a systematic review with meta-analysis. The Journal of Clinical Pharmacology, 62(4), 463–471.

Tedesco, S., Gajaram, G., Chida, S., Ahmad, A., Pentak, M., Kelada, M., … Soetan, O. T. (2021). The efficacy of MDMA (3, 4-methylenedioxymethamphetamine) for post-traumatic stress disorder in humans: a systematic review and meta-analysis. Cureus, 13(5).

TGA. (2023). Change to classification of psilocybin and MDMA to enable prescribing by authorised psychiatrists.

Thomas, J. L., Wilk, J. E., Riviere, L. A., McGurk, D., Castro, C. A., & Hoge, C. W. (2010). Prevalence of mental health problems and functional impairment among active component and National Guard soldiers 3 and 12 months following combat in Iraq. Archives of general psychiatry, 67(6), 614–623.

Ursano, R. J., Goldenberg, M., Zhang, L., Carlton, J., Fullerton, C. S., Li, H., … Benedek, D. (2010). Posttraumatic stress disorder and traumatic stress: from bench to bedside, from war to disaster. Ann N Y Acad Sci, 1208, 72–81. doi:10.1111/j.1749-6632.2010.05721.x

White, W. F., Burgess, A., Dalgleish, T., Halligan, S., Hiller, R., Oxley, A., … Meiser-Stedman, R. (2022). Prevalence of the dissociative subtype of post-traumatic stress disorder: a systematic review and meta-analysis. Psychol Med, 52(9), 1629–1644. doi:10.1017/s0033291722001647

Yehuda, R., Hoge, C. W., McFarlane, A. C., Vermetten, E., Lanius, R. A., Nievergelt, C. M., … Hyman, S. E. (2015). Post-traumatic stress disorder. Nature Reviews Disease Primers, 1(1), 1–22.

